# External Validation, Re-Calibration, and Extension of a Prediction Model of Early Acute Kidney Injury in Critically Ill Children using Multi-Center Data

**DOI:** 10.1101/2025.02.05.25321680

**Authors:** Adam C. Dziorny, Stephen Drury, Alex Clark, Reid WD Farris, Akira Nishisaki, Timothy T. Cornell, Daniel S. Tawfik, Tellen D. Bennett, Sareen S. Shah, Scott L. Weiss, Tahagod Mohamed, Neel Shah, James McMahon, Naveen Muthu, Randall C. Wetzel, Martin Zand, L. Nelson Sanchez-Pinto, PEDSnet and the PICU Data Collaborative

## Abstract

**Background:** Acute kidney injury (AKI) is common among children with critical illness and is associated with high morbidity and mortality. Risk prediction models designed for clinical decision support implementation offer an opportunity to identify and proactively mitigate AKI risks. Existing models have been primarily validated on single-center data, owing partly to the lack of appropriately detailed multicenter datasets.

**Objective:** To determine the accuracy of a single-center model to predict new AKI at 72 hours of ICU admission across two multicenter datasets and extend this model to improve prediction accuracy while maintaining acceptable alert burden.

**Derivation and Validation Cohorts:** We separately derived models in two datasets: PEDSNET-VPS, created through the linkage of PEDSnet electronic health record (EHR) extraction with Virtual Pediatric Systems (VPS); and the PICU Data Collaborative dataset, created through EHR extraction and harmonization from eight participating institutions. Derivation datasets comprised temporal and location-specific spit of these datasets (80%), while the holdout test split comprised the remaining (20%).

**Prediction Model:** We recalibrated an existing single-center model and measured discrimination and accuracy. We then add features guided by precision and recall measures. All features were available at 12 hours of ICU admission. We measure discrimination and accuracy at multiple cut-points and identify the features contributing most to the risk score.

**Results:** In two datasets comprising 186,540 ICU admissions, we report an incidence of early AKI of 2.2 – 2.7%. Initial recalibration of an existing single-center model demonstrated poor discrimination (AUROC 0.60 – 0.78). Following the addition of new features, we report higher AUROC values of 0.79 - 0.80 and AUPRC values of 0.13 – 0.21 in both datasets. We report accuracy at several cutpoints as well as cross-validate between datasets.

**Conclusions:** In this first use of two new multicenter datasets, we report improved discrimination and accuracy in a model designed specifically for implementation, balancing sensitivity and precision to predict patients at risk for AKI development.

## INTRODUCTION

Acute kidney injury (AKI) occurs in up to 25% of critically ill children and is associated with high morbidity and mortality.^1–3^ Critically, AKI is also more frequent among racial minorities and uninsured children.^4^ New-onset AKI within the first 72 hours of ICU admission, occurring in only 4% of ICU admissions in one single-center study, may be an important target for intervention.^5^ Despite the call for more automated and data-driven approaches to preventing AKI,^6^ its identification remains primarily reactive, based on serum creatinine and urine output, which are measures of established injury.^7^ As a result, we often miss the opportunity for early mitigation strategies including limiting nephrotoxic medications, fluid restriction, and early renal replacement therapies.^6,8^

Risk prediction models leveraging machine learning techniques coupled with user-centered clinical decision support (CDS) offer an opportunity to identify at-risk patients and intervene earlier.^9–11^ Existing AKI risk prediction models in critically ill children have been developed and validated primarily on single-center data.^5,12–14^ The Renal Angina Index (RAI) is an AKI risk scoring tool that has been evaluated in a multicenter trial,^15^ but it was developed using expert opinion instead of a data-driven approach, relies on intake and output data which may be inaccurate in the electronic health record (EHR), may represent already established injury in many cases, and in validation was noted to miss 18.9% of severe AKI cases.^16^ Similarly, a quality improvement program to screen nephrotoxic medication exposure demonstrated success when translated across institutions, but specifically excluded critically ill patients.^17,18^

The lack of appropriately detailed datasets poses a significant barrier to predictive model development and validation among children with critical illness. Quality improvement and administrative datasets such as Virtual Pediatric Systems and Pediatric Health Information System, while voluminous and standardized, are not sufficiently granular for model training for complex conditions like AKI.^19^ Additionally, these datasets are completed upon patient discharge, but when designing prediction models for implementation, one must examine only data available at the time the prediction will be made.^20^ Both the Medical Information Mart for Intensive Care III (MIMIC-III) ^21^ and the eICU Collaborative Research Database ^22^ are publicly-available datasets of critical care patients used to develop and validate ML algorithms and have been the basis for the development and validation of a multitude of AKI prediction models in adults,^23^ but no equivalent publicly-available dataset exists yet in pediatric critical care.

In this study, we leveraged two new granular, multi-institution datasets comprised of harmonized EHR data to validate and update an existing single-center model to predict early AKI. We updated and validated the existing model with a focus on prospective implementation to improve prediction accuracy while maintaining acceptable alert burden. Developing and externally validating this updated model is an important first step toward its prospective implementation to help clinicians reduce the burden AKI in this population through proven intervention and prevention strategies, such as nephrotoxin avoidance in high-risk patients.

## METHODS

### Study Population

We included two multi-institution datasets in this study. Both datasets are convenience samples of all children admitted to the pediatric intensive care units (PICUs) during the data collection period. In both datasets we excluded patients < 30 days of age and > 18 years of age.

The first (PEDSNET-VPS) was created through the probabilistic linkage of the Virtual Pediatric Systems (VPS, LLC, http://www.myvps.org) and the PEDSnet pediatric learning health system (PEDSNET).^24^ VPS is a quality improvement dataset comprised of all admissions into the pediatric intensive care unit at participating sites. Data are abstracted manually by trained abstractors. Limited laboratory and vital sign data are recorded; however, diagnoses and treatments are recorded reliably through manual chart review. PEDSNET is an EHR extracted and harmonized dataset following a standard common data model.^25^ We have previously described this probabilistic linkage validation using single-center deterministic linkage as the gold-standard.^26^ We applied the same linkage algorithm across six sites’ worth of data in both datasets. Briefly, site matches were identified by pair-wise matching patients from every site in VPS with patients from every site in PEDSNET and identifying the site pair with the lowest aggregate likelihood ratio (suggesting the most accurate matches). Using these site-wise pairs, we then matched patients between VPS and PEDSNET, applying the same likelihood ratio cutoff calculated in our single-center linkage. Individual sites chose their date range for submission, with most spanning >6 years and the most recent data being from 2017. We excluded one site without laboratory result times that were necessary to calculate hours from ICU admission.

The second dataset is comprised of an eight-site dataset from the PICU Data Collaborative (PDC).^27^ Briefly, this collaborative is an initiative among academic PICUs from across the United States to aggregate and harmonize granular EHR data for data science use cases. Each site contributes its own extracted and anonymized clinical data for patients admitted to their PICU using an agreed-upon data model. All date elements are converted into time difference measurements (e.g., seconds from hospital admission) and all local identifiers are removed. Data elements are harmonized centrally on an Azure cloud compute instance (Microsoft Azure, Redmond, WA) by content experts from each site. All computing occurs in Microsoft ML Studio within the same cloud compute instance. Individual sites chose the date range for their data submission spanning 2 to 11 years, with most sites submitting at least 8 years’ worth of data. While there are currently 19 member sites in the PDC, the dataset presented in this study includes only the first eight sites, with admissions up to and including 2022.

The institutional review board (IRB) for the University of Rochester determined use of the first dataset to be not human subjects research, and the second dataset as an exempt study. All procedures associated with this research were conducted in accordance with the ethical standards delineated in the Helsinki Declaration of 1975. We have followed the Transparent Reporting of a Multivariable Prediction Model for Individual Prognosis or Diagnosis (TRIPOD+AI) reporting guidelines for predictive models ^28^ as well as specialty-specific reporting guidelines.^29^

### Study Variables

We analyzed both datasets using the same approach. All features were available in both datasets, unless specifically noted. We collected demographic data including age at PICU admission and gender for all patients. We identified all laboratory measures resulted during the PICU admission and selected the 44 most common measures across the two datasets. We selected the most commonly recorded vital signs including fraction of inspired oxygen (FiO2), blood pressure (systolic [SBP], diastolic [DBP], and mean arterial <MAP>), oxyhemoglobin saturation (SpO2), heart rate (HR), respiratory rate (RR), temperature, weight and height. We identified diagnoses for immunocompromised conditions based on ICD-9 and ICD-10 codes.^30,31^ Several features were only available in the PEDSNET-VPS dataset including pre-admission cardiac arrest and admission for postoperative recovery, both of which were coded in the VPS dataset. Additionally, from the PEDSNET-VPS dataset we were able to reliably abstract medications and obtain counts of nephrotoxic medications by matching each medication name to its Anatomical Therapeutic Chemical (ATC) classification.^32^ Medication administration data were not available in the PDC dataset at the time of analysis. Neither dataset includes measures of geocoded socioeconomic status (e.g., Child Opportunity Index). We *a priori* chose not to include race or ethnicity as abstracted variables in our predictive model given the lack of biological plausibility related to race and ethnicity being associated with AKI, as opposed to social constructs potentially reflecting issues such as poor access to care and systemic racism, and to avoid including social determinants of health in a clinical decision-making use case that could exacerbate systemic bias.^33^

### Data Split

Each dataset was split into three cohorts, termed development, validation, and test, using temporal and site-specific sampling. In the PEDSNET-VPS dataset, the development and validation datasets were randomly split 80:20 and included all patients admitted before 06-01-2016, or approximately 75% of the dataset. The remaining 25% of patients admitted after 06-01-2016 were included in the test dataset. In the PDC datasets, the development dataset included all ICU admissions prior to 2019, the validation dataset included ICU admissions in 2019 – 2020, and the test dataset included ICU admissions in 2021 and beyond, as well as all admissions from one site (Supplementary Figure 1). Although datasets represent a convenience sample, we compared the sample sizes within each split to previous reports of early AKI incidence to ensure adequate cases for prediction.

### AKI Definition & Outcomes

We followed the Early AKI definition used in the original model by Sanchez-Pinto and Khemani, based on the Kidney Disease Improving Global Outcomes (KDIGO) staging criteria using serum creatinine (SCr).^3,5^ Briefly, we calculated baseline serum creatinine using a staged approach. We first attempted to identify the most recent SCr value within the prior 90 days. If that measure was unavailable, we calculated the height dependent SCr; if no height measure was available, we calculated the height-independent SCr.^34^ Outliers were removed based on exceeding two standard deviations from median.^35^ From these baseline SCr measures, we calculated the AKI stage at each time point a new SCr was obtained during the first three days of the hospitalization. Patients were excluded from subsequent analysis if they met criteria for AKI on ICU admission or within 12 hours following admission. Each remaining ICU admission was assigned the maximum stage AKI achieved between hour 12 and 72 following admission, from stage 0 (or no AKI) through stage 3. Patients with no measured creatinine values during this period were classified as stage 0 (no AKI). Neither dataset includes reliable intake and output measures, therefore we did not include measures of fluid balance or urine output-based AKI.

### Single Center Model Re-Calibration

We first measured the discrimination and accuracy of a previously described parsimonious single center model.^5^ We utilized the initial coefficients in the original publication, and then recalibrated the same original single-center model.^5^ Importantly, two of the seven variables (postoperative care and preadmission cardiac arrest) were not available in the PDC dataset. We did not impute or rescale any data for this validation; similar to the initial single center model, missing variables were simply not added to the score. Only variables available up to 12 hours following ICU admission were included in the model, choosing the value that resulted in the worse score if multiple values were available. We then re-calibrated and updated the coefficients by training a multivariable logistic regression model on our development data split in each dataset, and reporting results of the model on the test data split in each dataset. For this and subsequent models we computed area under the receiver operating characteristic curve (AUROC) and area under the precision recall curve (AUPRC), as this measure incorporates precision, or positive predictive value (PPV), and may be more informative for modeling problems with class imbalance.^36^ We report the sensitivity, specificity, and PPV at the 50^th^ and 90^th^ percentiles of the risk score in each dataset, and compare these to the validation cohort reported in the original study.^5^

### Additional Feature Selection and Model Training

To expand the single-center model based on published literature, we *a priori* defined additional physiologically plausible features for inclusion. Additional features include laboratory values, vital sign measurements, and presence of an immunocompromised diagnosis as described above. Again, we only included results and measurements available up to 12 hours following ICU admission. We aggregated repeated measures such as vital signs in several ways (e.g., maximum, minimum, mean, median) resulting in several candidate feature values for each measure. We measured pairwise collinearity between all feature pairs using the maximum of Pearson, Kendall, and Spearman correlation coefficients, and removed one feature from any pair where this coefficient was above 0.8. We encoded binary features, such as the presence of immunocompromised diagnosis, using a one-hot encoding approach (i.e. yes vs. no). All continuous features were scaled to minimize magnitude bias.

We performed feature selection simultaneous to imputation and model selection within the development split of each dataset. We imputed using either median imputation or median + last recorded value imputation, whereby the last recorded laboratory value was used instead of the median value. We compared targeting optimal AUROC vs AUPRC, and we compared four different model types: decision tree, random forest, penalized logistic regression, and support vector machines. We began the feature selection process with an initial feature set comprised of the seven features with the best performance as independent predictors of AKI. We then added one feature at a time to identify those which increased target measure performance. Then using backward elimination, we removed features if their absence did not decrease model performance. Finally, we trained each of the different model types on each feature set. Through this search space we were able to identify the optimal feature set, imputation method, and model type. Once we identified these optimal parameters, we performed final model hyperparameter tuning in the validation split of the datasets.

### Statistical Analysis

We performed initial dataset linkage (PEDSNET-VPS) and descriptive analysis in R version 4.2.1, including packages ‘dplyr’ and ‘ggplot’.^37,38^ Subsequent feature engineering, selection, model creation and analysis were completed in Python version 3.8 using packages ‘NumPy’, ‘pandas’, and ‘SciPi’.^39–41^

We assessed model accuracy in the test split of each dataset. We measured model discrimination as AUROC and AUPRC and calculated 95% confidence intervals (CI) using bootstrapping methodology. We tested differences between curves on the same dataset using DeLong’s test, however could not test significance between datasets as they are maintained on separate, secure resource compute environments.^42^ We calculated plot metrics including sensitivity, PPV, and number needed to alert (NNA, the inverse of PPV) across all potential cutpoints. We also assessed these metrics in the validation and test splits at each of the 50^th^ and 90^th^ percentile cutpoints, defined in the development split within each dataset. We cross-validated the final PDC model in the PEDSNET-VPS dataset and report the same metrics at the 50^th^ and 90^th^ percentile cutpoints. We report feature significance as contribution to the model based on Shapley Additive exPlanation (SHAP) scores. All source code for this project is available under the MIT license on GitHub (Microsoft Corporation, Redmond, WA): https://github.com/Lab-for-Integrated-Decision-Support/vps-peds-aki.

## RESULTS

### Epidemiology and Demographics

We identified a total of 67,167 ICU admissions across five sites in the PEDSNET-VPS dataset. Similarly, we identified a total of 154,174 ICU admissions across eight sites in the PDC datasets. After excluding admissions based on patient age, the length of ICU stay < 12 hours, and AKI on admission or within the first 12 hours, there were 55,890 (83.2%) and 130,650 (84.7%) eligible ICU admissions, respectively. The incidence of new AKI by 72 hours was 2.2% (1,240 admissions) and 2.7% (3,535 admissions) in each dataset, with >60% of these in both datasets being Stage 1 AKI. The PDC dataset contained patients who were generally younger on admission, had longer median ICU length of stay, and higher mortality (Table 1).

**Table 1.** Demographic features of the two cohorts of ICU admissions.

|  | **PEDSNET-VPS** | **PDC** |
| --- | --- | --- |
| All ICU Encounters | 82,457 | 154,174 |
| Excluded Encounters |  |  |
| Incomplete Lab Result Times <sup>a</sup> | 15,290 (18.5%) | -- |
| ICU Length of Stay < 12 hours <sup>a</sup> | 7,837 (11.7%) | 15,454 (10.0%) |
| AKI < 12 hours after ICU Admission <sup>a</sup> | 3,440 (5.7%) | 8,070 (5.8%) |
| ICU Encounters meeting inclusion<br>(% of all ICU encounters) | 55,890 | 130,650 |
| Unique Patients | 34,686 | 86,202 |
| Early AKI by 72 hours (%) | 1,240 (2.2%) | 3,535 (2.7%) |
| KDIGO AKI Stage 1 (%) | 762 (1.4%) | 2,927 (2.2%) |
| KDIGO AKI Stage 2 or 3 (%) | 478 (0.8%) | 630 (0.5%) |
| Age, median years (IQR) | 4.2 (1.2 – 10.9) | 3.2 (0.7 – 10.2) |
| Gender, Female (%) | 24,679 (44) | 58,247 (45) |
| PICU LOS, median days (IQR) | 1.6 (0.8 – 3.5) | 2.0 (1.1 – 4.7) |
| Hospital Mortality (% of hospital encounters) | 708 (1.3%) | 2,002 (2.1%) |
| Cohort split |  |  |
| Development | 33,300 (60%) | 76,762 (59%) |
| Validation | 8,326 (15%) | 27,132 (21%) |
| Test | 14,264 (20%) | 26,756 (20%) |

<sup>a</sup> Percentage is calculated as a percent of the remaining encounters after subtracting the prior exclusions. For example, the percentage of PEDSNET-VPS encounters with “ICU Length of Stay < 12 hours” is as a percentage of those eligible after excluding those encounters with “Incomplete Lab Result Times.”

### Single-Center Model Performance

We applied existing coefficients to the original Early AKI prediction model in both datasets and found moderate-to-poor discrimination (AUROC: PEDSNET-VPS 0.74, PDC 0.39). After re-calibration in the development split, we found modestly improved performance in both datasets (AUROC: PEDSNET-VPS 0.78 [95% CI: 0.78 – 0.78], PDC 0.65 [95% CI: 0.64 – 0.65]). Similarly, at both the 50^th^ and 90^th^ percentile cutpoints, sensitivity and PPV were lower than the original single-center model’s validation dataset (Table 2).

**Table 2.** Discrimination and accuracy of the single-center model recalibrated on the development split of each dataset. Results are reported from the test split in each dataset.

|  | **Temporal Validation**<br>**Sanchez-Pinto et al <sup>a</sup>** |  | **PEDSNET-VPS** |  | **PDC <sup>b</sup>** |  |
| --- | --- | --- | --- | --- | --- | --- |
| Total N | 2,832 |  | 14,264 |  | 26,756 |  |
| Early AKI (%) | 114 (4) |  | 303 (2) |  | 574 (2) |  |
| AUROC (95% CI) | 0.86 (0.85 – 0.88) |  | 0.78 (0.78-0.78) |  | 0.65 (0.64-0.65) |  |
| AUPRC (95% CI) | Not reported |  | 0.12 (0.12-0.12) |  | 0.10 (0.10-0.11) |  |
| Cutpoint Percentile | 50<sup>th</sup> | 90<sup>th</sup> | 50<sup>th</sup> | 90<sup>th</sup> | 50<sup>th</sup> | 90<sup>th</sup> |
| Test Positive at Cutpoint (%) | 1578 (56) | 493 (17) | 7134 (50) | 1427 (10) | 13277 (50) | 2723 (10) |
| Sensitivity % (95% CI) | 96 (90-98) | 71 (62-79) | 85 (81-89) | 37 (32-42) | 66 (63-70) | 29 (25-32) |
| Specificity % (95% CI) | 50 (50-50) | 86 (86-86) | 51 (50-52) | 91 (90-91) | 50 (50-51) | 90 (90-91) |
| PPV % (95% CI) | 7 (7-7) | 16 (14-18) | 4 (3-4) | 8 (6-9) | 3 (3-3) | 6 (5-7) |

<sup>a</sup> For comparison we include results from the Validation #2 dataset from the original publication, Sanchez-Pinto et al, 2016
<sup>b</sup> Two of the seven features from the single-center model (pre-operative status and cardiac arrest prior to admission) were unavailable in this dataset.

### Expanded Model Derivation and Performance

We identified 45 laboratory tests for possible inclusion, along with 10 vital signs (Supplementary Table 1). Imputation and target metric search identified last value + median imputation targeting AUPRC to result in the highest accuracy measures (Supplementary Table 2). We chose logistic regression for the final model as it provided an optimal combination of metric accuracy and explainability. In the test splits, AUROC were 0.80 in PEDSNET-VPS and 0.82 (95% CI: 0.82 – 0.82) in PDC. On the other hand, the AUPRC was higher in the PDC dataset (0.25) compared to PEDSNET-VPS (0.13) (Figure 1). Accordingly, at the 90^th^ percentile cutpoint, PPV was higher in the PDC dataset (18) compared to the PEDSNET-VPS dataset (10) (Figure 2). Sensitivity and PPV were lower at both cutpoints in the cross-validated PEDSNET-VPS dataset when applying the PDC model (Table 3). Features most contributing to the prediction score included calcium, lactate, creatinine, and systolic blood pressure (Supplementary Table 3).

**Figure 1.**
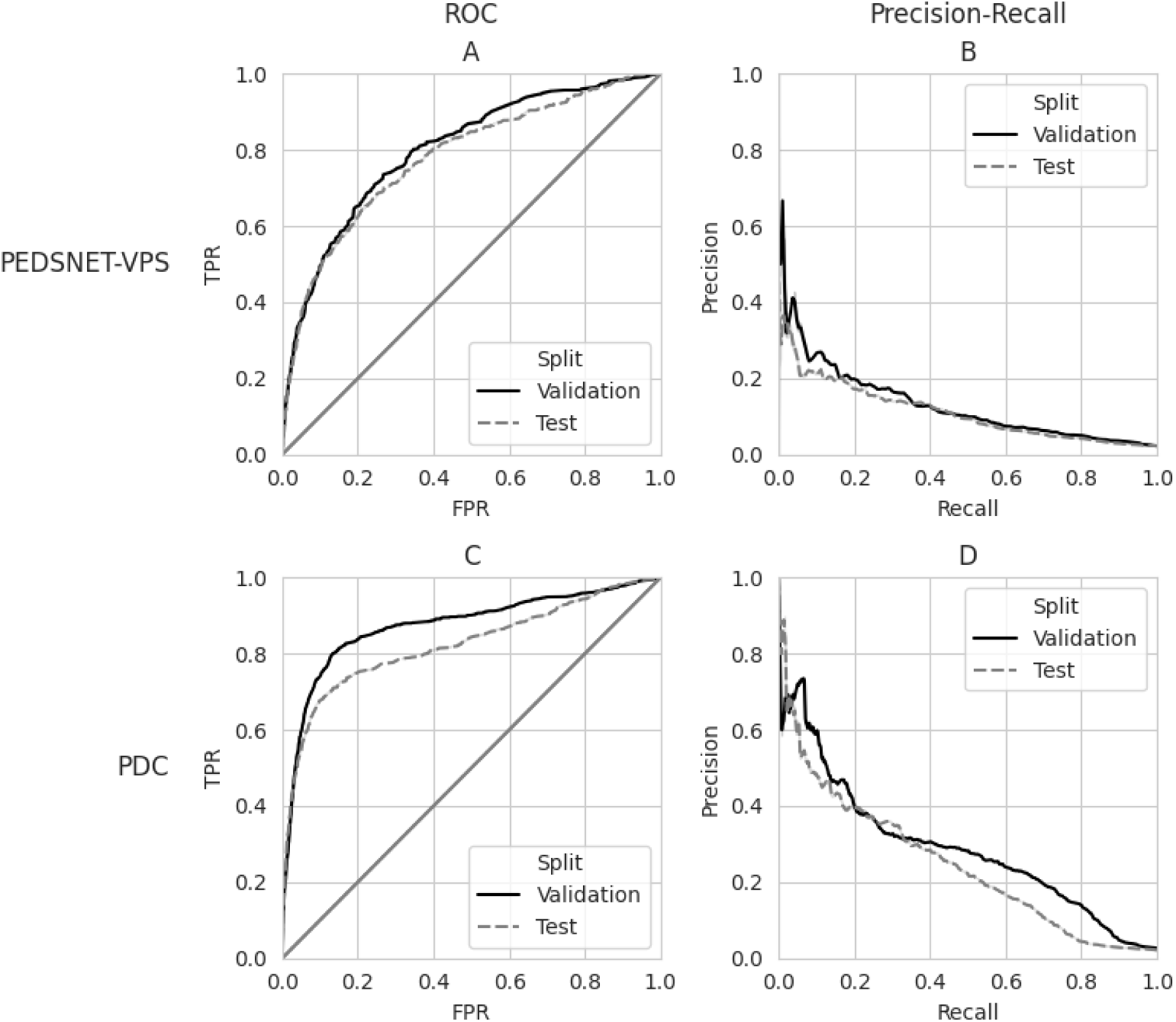
AUPRC and AUROC curves of best-performing logistic regression model following the addition of new features. Columns represent the Receiver Operating Characteristic (ROC) curves (A, C) and Precision-Recall curves (B, D) for each dataset. Rows represent the PEDSNET-VPS dataset (A, B) and the PDC dataset (C, D). All curves report data from the validation (solid line) and test (dashed line) splits.

**Figure 2.**
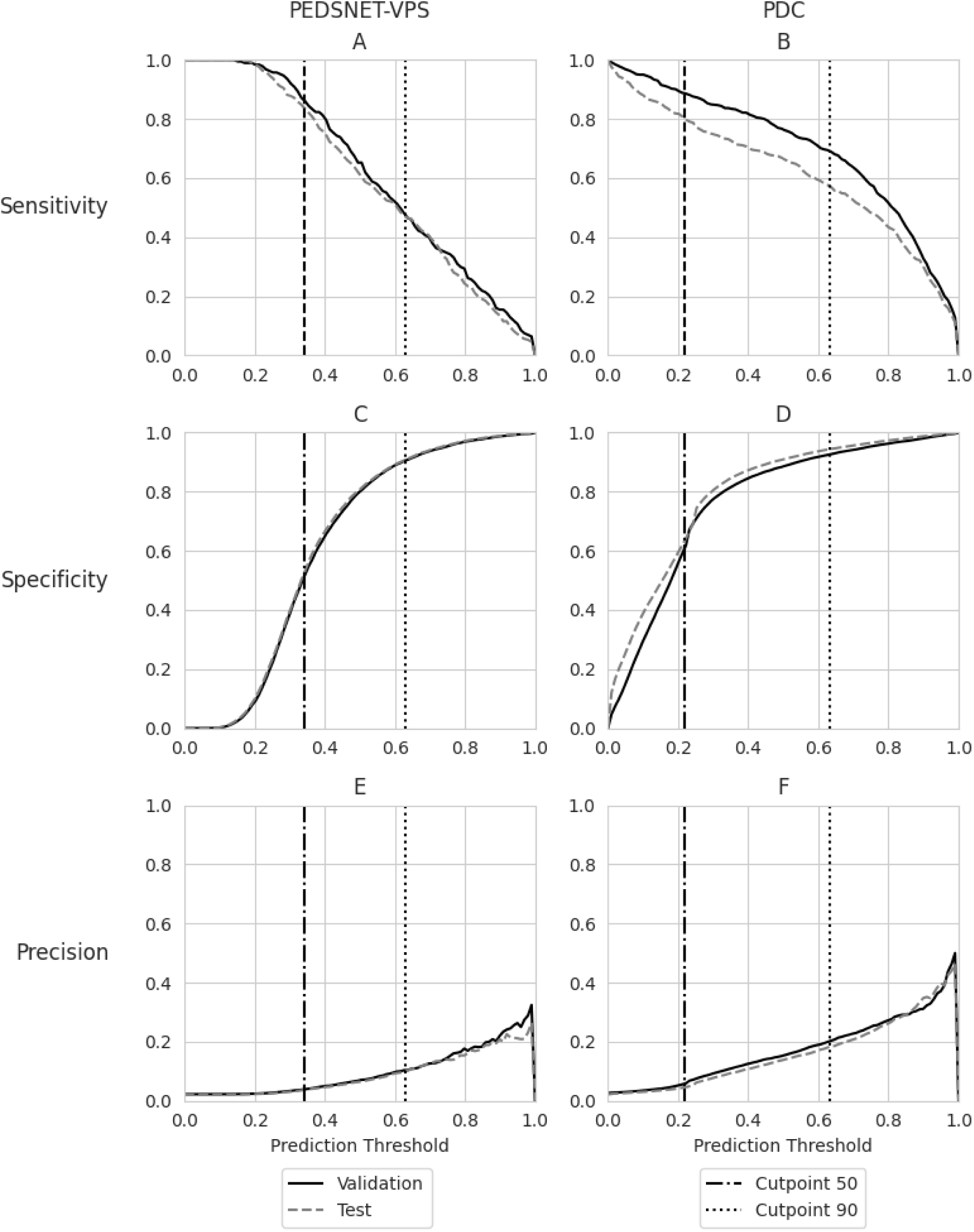
Sensitivity, specificity, and precision across the range of cutpoints for the best-performing logistic regression model in each dataset, reported from the test split. Datasets are indicated for each column. In each dataset, the 50^th^ and 90^th^ percentile cutpoints are indicated by vertical lines.

**Table 3.** Final model diagnostic performance for the optimal (“best”) model with the optimal feature set, including only those features able to be extracted from the EHR using FHIR-based calls within the first 12 hours of ICU admission.

|  | PEDSNET-VPS |  |  |  | PDC |  |  |  |  |  |
| --- | --- | --- | --- | --- | --- | --- | --- | --- | --- | --- |
|  | Val |  | Test |  | Val |  | Test |  | Test 2<br>(PEDSNET-VPS) |  |
| Total N | 8,326 |  | 14,264 |  | 27,132 |  | 26,756 |  | 14,264 |  |
| AUROC<br>(95% CI) <sup>c</sup> | 0.80 |  | 0.80 |  | 0.88<br>(0.88 – 0.88) |  | 0.82<br>(0.82 – 0.82) |  | 0.68<br>(0.68 – 0.68) |  |
| AUPRC<br>(95% CI) <sup>c</sup> | 0.13 |  | 0.13 |  | 0.30<br>(0.30 – 0.31) |  | 0.25<br>(0.24 – 0.25) |  | 0.09<br>(0.09 – 0.09) |  |
| Cutpoint<br>Percentile <sup>a</sup> | 50<sup>th</sup> | 90<sup>th</sup> | 50<sup>th</sup> | 90<sup>th</sup> | 50<sup>th</sup> | 90<sup>th</sup> | 50<sup>th</sup> | 90<sup>th</sup> | 50<sup>th</sup> | 90<sup>th</sup> |
| Risk test<br>positive (%) | 4135<br>(51) | 854<br>(10) | 6,924<br>(49) | 1,418<br>(10) | 11,126<br>(41) | 2,450<br>(9) | 10,149<br>(38) | 1,798<br>(7) | 1,580<br>(11) | 657<br>(5) |
| Sensitivity | 87 | 48 | 83 | 47 | 89 | 69 | 80 | 57 | 44 | 30 |
| Specificity | 51 | 91 | 52 | 91 | 60 | 93 | 63 | 94 | 90 | 96 |
| PPV | 4 | 10 | 4 | 10 | 6 | 20 | 5 | 18 | 8 | 14 |
| NNA <sup>b</sup> | 26 | 11 | 26 | 11 | 18 | 5 | 23 | 6 | 12 | 8 |

<sup>a</sup> For each model (PEDSNET-VPS and PDC), cutpoint percentiles were established on development split of each dataset and applied to additional datasets. For PDC model, cutpoints from development split were applied to PDC Test split as well as PEDSNET-VPS Test split.
<sup>b</sup> NNA = Number Needed to Alert, or 1 / Precision
<sup>c</sup> Confidence intervals were calculated by 10-fold sampling (with replacement) of 3000 samples, followed by sorting results and calculating the 0.025 and 0.975 percentiles.

## DISCUSSION

We report the first use of two new multicenter pediatric critical care datasets to evaluate, re-calibrate, extend, and validate a machine learning-based prediction algorithm of new early AKI. We first applied an existing single-center model of early AKI prediction across two multi-institutional datasets and demonstrate poor discrimination and accuracy compared to the initial single-center validation. After the addition of new features, we report improved discrimination and accuracy using features that would be available at the time of the prediction. The development of the new model focused on implementability, selecting features commonly available at 12 hours after admission and targeting metrics balancing both sensitivity and PPV.

Several other risk prediction algorithms have been developed to identify AKI, but each have limitations.^11^ Sanchez-Pinto and Khemani created a parsimonious score to predict early acute kidney injury from single-center data, however when we applied this score across multicenter data the accuracy decreased.^5^ Additionally, this algorithm relies on data which may not be stored in discrete variables within the EHR (e.g., pre-admission cardiac arrest). Similarly, Wang and colleagues developed a single-center derived model to predict AKI risk throughout the ICU stay. They report much higher proportion of AKI (470 / 866, or 54%) in their validation cohort as they included cases beyond the first 72 hours of the ICU admission and reported a lower AUROC of 0.74 (95% CI 0.71 – 0.77) in the ICU cohort. Dong et al. created a model based on three independent centers’ EHR data using engineered features to predict AKI development within 30 hours.^14^ While they report excellent discrimination (AUROC 0.89) and an impressive PPV (0.47), this result is misleading as it would apply to actual real-time implementation. They focused on *encounter-level analysis* of patients who did and did not develop AKI, such that control encounters (e.g., those were no AKI was observed) were only counted once. To provide an accurate measure of implemented PPV, and thus alert burden, authors must account for this model running in real-time across every analysis window for every admitted patient, which would substantially increase the number of possible true and false negatives.

In addition to the above machine learning-based prediction algorithms, risk scores have been developed and validated in general PICU patients as well as in specific subpopulations. The Renal Angina Index (RAI) combines patient risk factors and potential renal injury early in the PICU admission to identify patients at risk of developing or having persistent AKI three days after PICU admission (e.g., from 72 – 96 hrs). The score incorporates SCr measurement and percent fluid overload as well as features of critical illness (e.g., mechanical ventilation and inotropy use). Initial validation studies included patients with AKI on admission as well as patients with sepsis and thus higher rates of early AKI (10.2 – 19.4%).^15^ A follow-up validation study from prospectively obtained multicenter data also included sicker patients with higher mortality (5.1%) and higher rates of severe AKI by ICU day 3 (23.1%).^16^ In this population, less reflective of the general ICU admissions, while RAI demonstrated a reasonable PPV (121 / 286, or 42%), sensitivity was poor (121 / 368, or 32%) and 18.9% of patients with severe AKI were missed. Incorporating biomarker data and limiting to specific populations with sepsis results in improved performance, but may not be useful when considering all ICU admissions.^43,44^

Multicenter validation of single-center predictive models is critical for generalization outside of the original development site. Since AKI occurs across the sociodemographic spectrum, prediction solutions should be generalizable and shareable to cross the “digital divide” and improve care equity.^45^ As we show here, and others have shown, single-center models often do not show adequate performance in external validation, even with re-calibration.^46^ Rockenschaub et al. compared single-center derived models across two adult ICU EHR-derived datasets and even the addition of newer generalizability criterion did not improve performance substantially.^46^ Unfortunately, we are limited by the availability of sufficiently complete, harmonized datasets in pediatric critical care to perform this kind of external validation.^19^ We anticipate that efforts like the PICU Data Collaborative may serve an important role in development, recalibration, and validation of generalizable models.^27^ Transportability, or adequate performance across multiple sites, is critical for trial design and testing as well as implementation in new patient populations, specifically at sites without informatics expertise such as potentially less-well-resourced institutions.^47^

We intentionally focused model development and feature selection to design for implementation. Only a very small percentage of published models are translated into the clinical environment through real-time clinical decision support (CDS).^29^ As highlighted in the recent Phoenix Criteria for Sepsis and Septic Shock in Children study, the AUPRC is more informative than the AUROC for imbalanced problem (i.e., with rare events of interest) and in acute care use cases such as ours, where sensitivity and PPV provide more actionable insights (e.g. NNA, false alert burden, missed cases, etc.).^36^ We report PPV and NNA in addition to sensitivity as these measures are important to clinicians who may be receiving these alerts. We chose the logistic regression model as this is more interpretable for treating clinicians, compared to SVM or random forest models.

We are implementing our current model using a Fast Healthcare Interoperability Resource (FHIR) based interface and user-focused design to improve the actionability and usability of this model.^48^ The addition of urine biomarkers in a tiered approach, similar to the TAKING FOCUS 2 trial, may allow for acceptance of higher rates of false positives to enrich the population for subsequent biomarker stratification and intervention (e.g., furosemide stress test) assessments.^49^ While CDS-based prediction models may improve screening rates,^50,51^ not all early identification improves clinical outcomes.^52^ Future studies should incorporate pragmatic clinical trial design across multiple institutions, using generalized, shareable, user-designed design support, possibly incorporating biomarkers and protocolized care bundles.

This study had several limitations to highlight. First, one of our datasets (PEDSNET-VPS) included a large proportion of excluded subjects due to missing data and did not include data more recent than 2017. Neither dataset currently includes intake or output (IO) fluid measurements and therefore we could not compute AKI based on all KDIGO criteria, nor could we compare the model to the RAI score. Both datasets currently lack harmonized medication data to accurately report continuous infusions of vasoactive medications as well as harmonized mechanical ventilation data. There was overlap among two of the sites in these multi-institution datasets, however with the reduced time window (e.g., up to 2017 data) in the PEDSNET-VPS dataset we did not exclude these overlapped sites. We anticipate that with continued addition of new data into the PICU Data Collaborative dataset, including IO fluid measurements, harmonized medications, and harmonized mechanical ventilation, we will be able to update, recalibrate, and revalidate our model.

## CONCLUSION

In the first use of two new multicenter datasets specifically designed for machine learning development and validation, we demonstrate improved discrimination and accuracy over re-calibration of a single-center model. We designed our model and selected features for implementation, balancing sensitivity and PPV to both capture patients at risk for new early AKI while balancing alert burden to clinicians. Ongoing work includes prospective silent validation in our intensive care environment as well as continuous model improvement through the addition of new temporal and site-specific data.

## Data Availability

All software and work product produced in the present study are available upon reasonable request to the authors. Data from the two contributing datasets [PEDSNET-VPS and PDC] are used under agreement from the controlling entities and the authors cannot grant requests for access to these data.

## ACKNOWLEDGEMENTS

The authors would like to thank all those who supported data collection, aggregation, and harmonization at each site. We would like to thank the Virtual PICU participating centers and research coordinators, as well as the PEDSnet data coordinating center analysts, programmers, and administrative support personnel. Additionally, we would like to thank the PICU Data Collaborative project manager Jeffrey Terry and data team members including Alysia Flynn, Ruiqi Huang, Eugene Laksana, Ishmael Obeso, Michael Reilly, Alexsandra Kretsu and Long Ho.

## KEY POINTS

### Question

- This study aims to leverage two new granular, multi-institution datasets to validate and update an existing single-center model to predict early AKI, with a focus on prospective implementation, to improve prediction accuracy while maintaining acceptable alert burden.

### Findings

- We validated an existing single-center model in two multi-institution datasets with 55,890 and 130,650 ICU admissions, respectively. Applying existing model coefficients resulted in moderate-to-poor discrimination in both datasets, which improved slightly (AUROC: 0.65 - 0.78) by recalibrating using the same feature set. Following the iterative addition and removal of contributing features, we improved discrimination in both datasets (AUROC: 0.80 – 0.82, AUPRC: 0.13 – 0.25). Sensitivity and precision across multiple cutpoints in the test data split were similar or improved compared to existing single center models.

### Meaning

- Machine learning models updated in granular, multi-institution datasets may help predict new early AKI when implemented prospectively. Models may be more transportable to sites with new patients and decreased informatics expertise. Implemented models may help clinicians reduce the burden of AKI through intervention and prevention strategies.

## SUPPLEMENTARY TABLES AND FIGURES

**Supplemental Figure 1.**
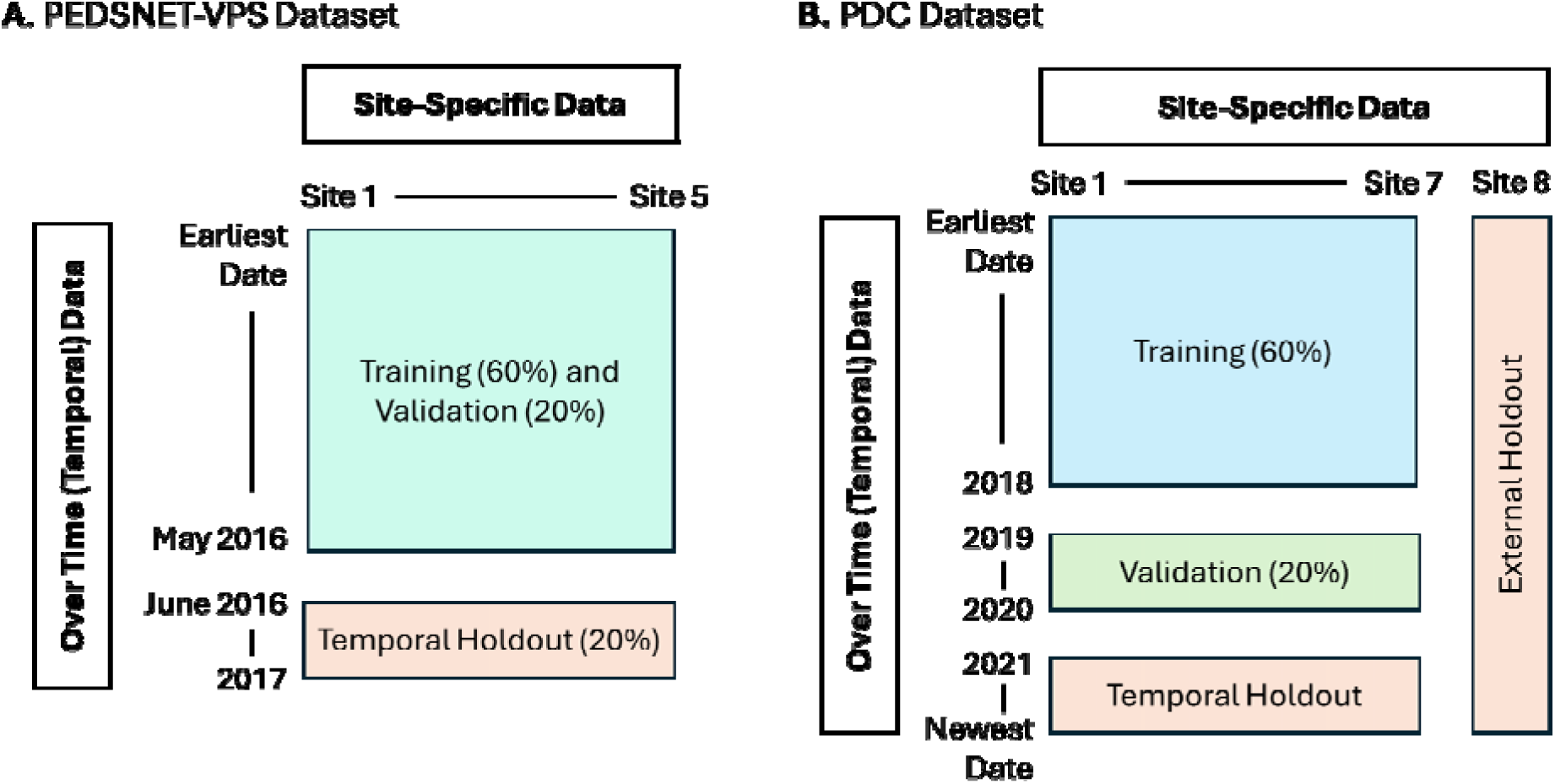
Schematic of data splits in each of the two datasets.

**Supplemental Table 1.**
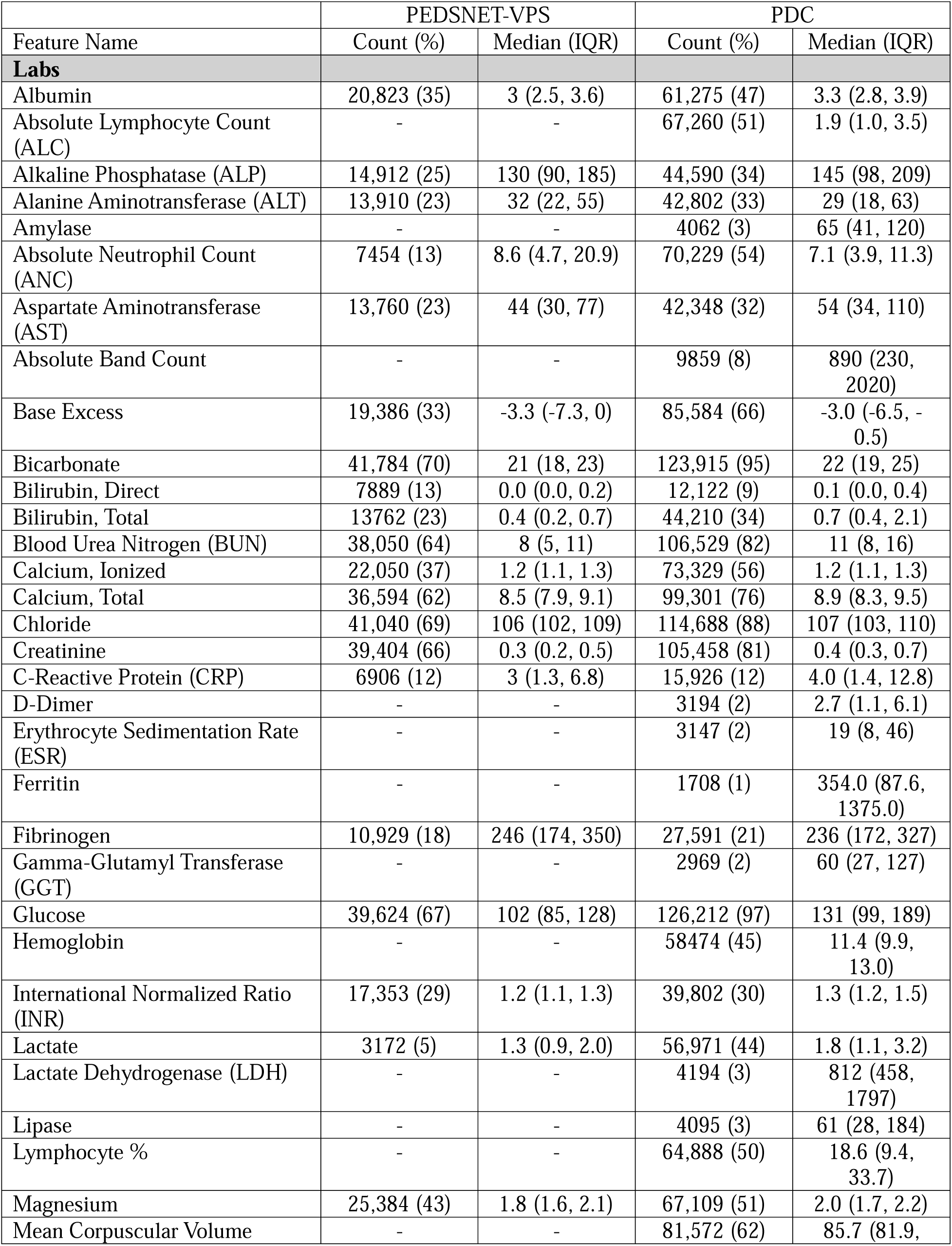
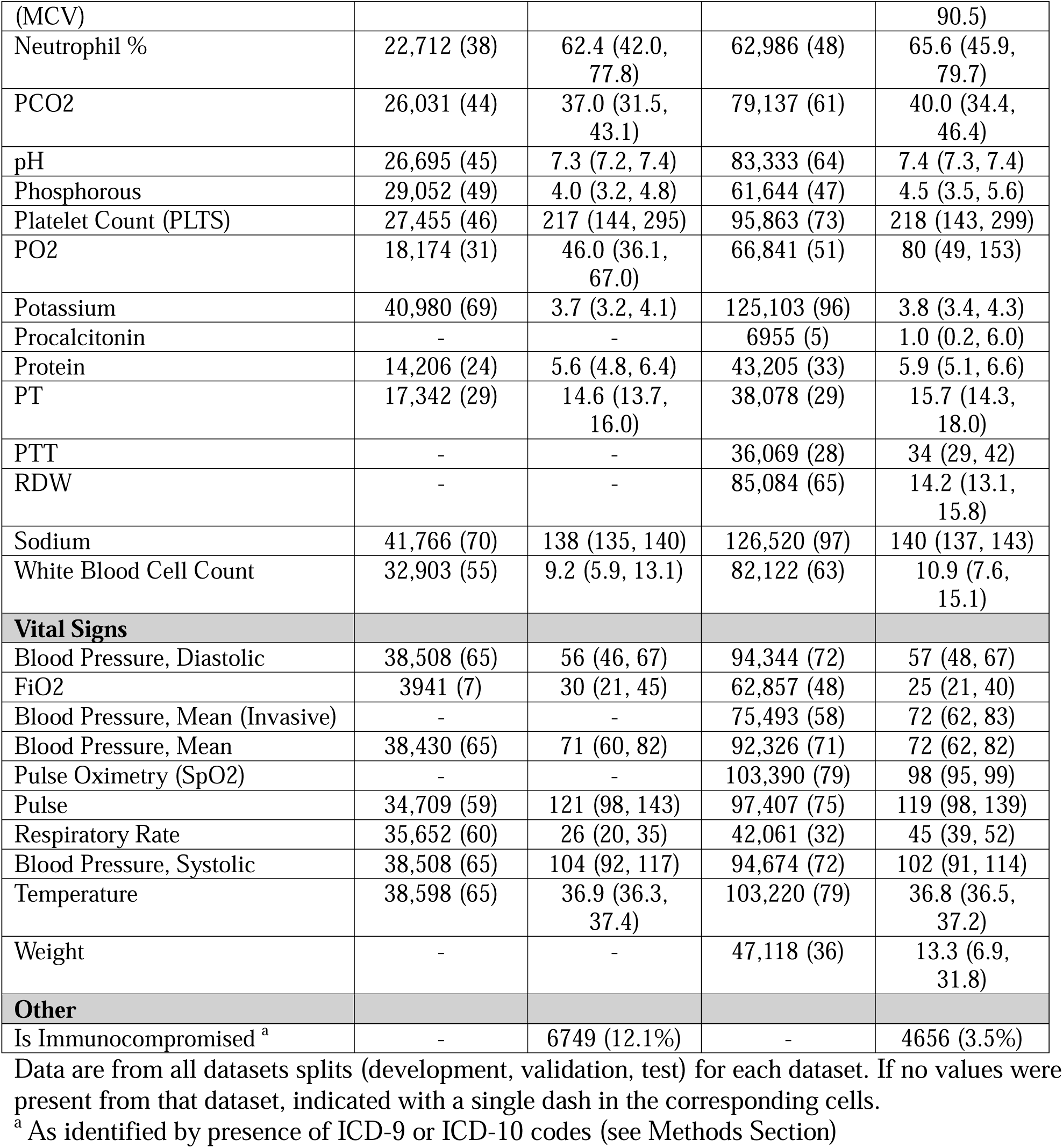
Summary of feature counts and median values for all features considered for model extension.

**Supplementary Table 2.** Feature selection imputation and model optimization results.

| **PEDSNET-VPS** |  |  |  |  |  |  |  |  |  |
| --- | --- | --- | --- | --- | --- | --- | --- | --- | --- |
|  |  | **AUROC Targeted** |  |  |  | **AUPRC Targeted** |  |  |  |
|  | **Model** | **AUROC** | **Sens** | **Spec** | **PPV** | **AUPRC** | **Sens** | **Spec** | **PPV** |
| **Median Impute** | Decision Tree | 0.54 | 10 | 98 | 9 | 0.03 | 13 | 98 | 11 |
|  | Random Forest | 0.76 | 86 | 39 | 3 | 0.12 | 84 | 51 | 4 |
|  | Logistic Regression | 0.79 | 85 | 51 | 4 | 0.14 | 86 | 51 | 4 |
|  | SVM | 0.80 | 87 | 51 | 4 | 0.14 | 89 | 51 | 4 |
| **Median + Last Impute** | Decision Tree | 0.56 | 14 | 98 | 13 | 0.03 | 11 | 98 | 11 |
|  | Random Forest | 0.78 | 91 | 41 | 3 | 0.12 | 89 | 44 | 4 |
|  | Logistic Regression | 0.82 | 90 | 51 | 4 | 0.15 | 89 | 44 | 4 |
|  | SVM | 0.82 | 90 | 51 | 4 | 0.15 | 90 | 51 | 4 |
| **PDC** |  |  |  |  |  |  |  |  |  |
|  |  | **AUROC Targeted** |  |  |  | **AUPRC Targeted** |  |  |  |
|  | **Model** | **AUROC** | **Sens** | **Spec** | **PPV** | **AUPRC** | **Sens** | **Spec** | **PPV** |
| **Median Impute** | Decision Tree | *0.58 | 19 | 98 | 18 | 0.05* | 18 | 98 | 17 |
|  | Random Forest | 0.86 | 93 | 45 | 4 | 0.26 | 93 | 45 | 4 |
|  | Logistic Regression | 0.79 | 83 | 51 | 4 | 0.18 | 84 | 51 | 4 |
|  | SVM<sup>a</sup> | 0.86 | 92 | 51 | 5 | 0.21 | 94 | 51 | 5 |
| **Median + Last Impute** | Decision Tree | *0.60 | 22 | 98 | 20 | 0.07* | 23 | 98 | 22 |
|  | Random Forest | 0.91 | 96 | 49 | 5 | 0.33 | 96 | 50 | 5 |
|  | Logistic Regression | 0.86 | 89 | 51 | 5 | 0.29 | 90 | 51 | 5 |
|  | SVM | 0.87 | 91 | 48 | 5 | 0.17 | 94 | 51 | 5 |

Discrimination and accuracy results for feature selection within each dataset, by imputation type, by feature selection target (AUROC vs AUPRC), and by model type. Accuracy results (sensitivity, specificity, and precision) are shown for the 50<sup>th</sup> percentile cutpoint in the validation split of each dataset. Sens = Sensitivity; Spec = Specificity; PPV = Positive Predictive Value, equivalent to Precision; SVM = Support Vector Machine.
<sup>a</sup> SVM used the radial basis function kernel

**Supplementary Table 3.** SHApley Additive exPlanation (SHAP) values for each included covariate in the final logistic regression PDC model.

| Covariate Name | SHAP Value |
| --- | --- |
| CALCIUM_ION_max | 1.82E-01 |
| CALCIUM_ION_min | 1.22E-01 |
| CALCIUM_ION_mean | 1.18E-01 |
| CREATININE_median | 4.07E-02 |
| CREATININE_mean | 4.01E-02 |
| LACTATE_mean | 2.88E-02 |
| SBP_mean | 2.42E-02 |
| MCV_mean | 2.03E-02 |
| BUN_mean | 1.69E-02 |
| CHLORIDE_max | 1.58E-02 |
| PLTS_mean | 1.29E-02 |
| BICARBONATE_min | 1.22E-02 |
| PH_min | 1.12E-02 |
| MAGNESIUM_min | 1.11E-02 |
| PROTEIN_max | 1.05E-02 |
| NEUTRO_PCT_mean | 1.03E-02 |
| BICARBONATE_max | 1.01E-02 |
| BASE_EXC_min | 9.06E-03 |
| RESP_RATE_mean | 8.79E-03 |
| GLUCOSE_min | 8.74E-03 |
| BILIRUBIN_TOT_max | 8.72E-03 |
| FIO2_max | 7.30E-03 |
| BILIRUBIN_DIR_max | 7.13E-03 |
| MAP_mean | 7.06E-03 |
| PROTEIN_min | 6.68E-03 |
| BASE_EXC_mean | 6.23E-03 |
| PCO2_max | 6.19E-03 |
| CRP_mean | 5.70E-03 |
| SPO2_min | 5.30E-03 |
| CHLORIDE_mean | 4.65E-03 |
| SODIUM_mean | 4.32E-03 |
| RDW_mean | 4.24E-03 |
| ALC_min | 3.69E-03 |
| PULSE_mean | 3.20E-03 |
| PT_min | 2.73E-03 |
| DBP_max | 2.41E-03 |
| SBP_min | 1.93E-03 |
| BILIRUBIN_DIR_median | 1.78E-03 |
| POTASSIUM_max | 1.77E-03 |
| ANC_mean | 1.70E-03 |
| ESR_min | 1.68E-03 |
| ESR_median | 1.67E-03 |
| PHOSPHORUS_min | 1.39E-03 |
| WEIGHT_mean | 1.38E-03 |
| TEMP_min | 1.30E-03 |
| AST_mean | 9.84E-04 |
| PCO2_mean | 9.04E-04 |
| BILIRUBIN_DIR_min | 8.53E-04 |
| PO2_max | 8.04E-04 |
| BASE_EXC_median | 4.20E-04 |
| CHLORIDE_min | 3.84E-04 |
| SBP_median | 1.83E-04 |
| PT_median | 7.77E-05 |
| ALBUMIN_max | 6.14E-05 |
| BANDS_ABS_mean | 0.00E+00 |

